# Statistical Applications in Pediatric Endocrinology: A Simulation on Metformin’s Effect on HbA_1*c*_ in High-Risk Adolescents

**DOI:** 10.1101/2025.10.12.25337852

**Authors:** Wei Shi

## Abstract

Metformin has been increasingly used off-label in adolescents with type 1 diabetes (T1D) or prediabetic conditions to improve glycemic control. Despite clinical trials suggesting modest improvements in HbA_1*c*_, evidence remains mixed and methodologically limited. This paper introduces a simulation framework to evaluate advanced causal inference estimators—including Targeted Maximum Likelihood Estimation (TMLE), Double Machine Learning (DML), and Bayesian Causal Forests (BCF)—for estimating the causal effect of metformin on HbA_1*c*_ reduction in high-risk adolescents. The framework incorporates realistic confounding structures based on empirical data and provides theoretical derivations for estimator properties.

## 1 Introduction

The management of type 1 diabetes (T1D) in adolescents presents a significant clinical challenge, compounded by the rising prevalence of insulin resistance and obesity in this population (Maahs et al., 2010). Poor glycemic control during these formative years can lead to long-term complications, making the search for effective adjunct therapies a clinical priority. Metformin, a biguanide that improves insulin sensitivity, has been widely investigated as an adjunct to insulin therapy. However, the evidence from randomized controlled trials (RCTs) has been notably inconsistent.

While some studies suggest modest, short-term improvements in glycated hemoglobin (HbA_1*c*_) (Libman et al., 2015), others, including large-scale meta-analyses, have found no sustained benefits, questioning its widespread use in this context (Abdel-Moniem et al., 2021). This evidentiary conflict highlights a fundamental methodological challenge: the complex interplay of metabolic, behavioral, and clinical factors that confound treatment effects. Observational studies, which reflect real-world clinical practice, are particularly susceptible to biases that traditional regression models fail to adequately address (Hernán, 2018).

This issue is part of a broader movement in biomedical research toward more rigorous statistical methods, especially in large-scale settings where controlling for false discoveries is paramount (Benjamini and Hochberg, 1995; Storey, 2002; Stephens, 2017). Indeed, the development of novel statistical frameworks, often drawing from empirical Bayes principles, has been crucial for making sense of high-dimensional data in fields like genomics and proteomics (Efron et al., 2001; Efron, 2007). For example, new hypothesis testing strategies have been specifically designed to handle the complex variance structures found in peptide array data (Zheng et al., 2021).

The successful application of advanced statistical models in other areas of biomedical science provides a roadmap. In immunology, for instance, sophisticated modeling has been essential for dissecting the complex epitope landscape of autoantibodies in diseases like rheumatoid arthritis, Sjögren disease, and even post-viral syndromes (Zheng et al., 2020; Mergaert et al., 2022; Amjadi et al., 2024; Parker et al., 2024; Bashar et al., 2024).

Similarly, the development of robust statistical techniques to correct for non-random assignment in experiments has enhanced the reliability of A/B testing (Zheng and Liu, 2024). The broad utility of these rigorous quantitative approaches is further demonstrated by their application in diverse fields, including cognitive science (Liu et al., 2022).

Inspired by these successes, this paper argues that modern causal inference methods offer a powerful solution to the metformin problem. We focus on estimators that are doubly robust and can accommodate complex data, such as Targeted Maximum Likelihood Estimation (TMLE) (van der Laan and Rose, 2011), Double Machine Learning (DML) (Chernozhukov et al., 2018), and Bayesian Causal Forests (BCF) (Hahn et al., 2020).

To demonstrate their value, we develop a comprehensive simulation environment where the ground-truth causal effect of metformin on HbA_1*c*_ is known. This framework allows for a direct and unbiased comparison of these advanced estimators, providing a clear illustration of their potential to generate more reliable evidence for clinical decision-making in pediatric endocrinology.

## 2 Simulation Framework

Let *A* denote treatment assignment (*A* = 1 for metformin, *A* = 0 for control), *Y* the post-treatment HbA_1*c*_, and *X* a vector of baseline covariates (e.g., age, BMI *z*-score, insulin dose, baseline HbA_1*c*_). The data-generating process is defined as:

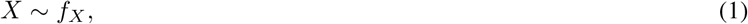

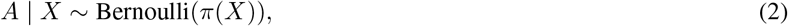

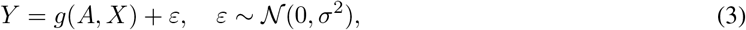

where *g*(*A, X*) incorporates nonlinear and interaction effects.

### 2.1 Structural Model

To simulate realistic clinical scenarios, we specify a structural model linking baseline covariates, treatment assignment, and outcome. Let *X* = (*X*_1_, *X*_2_, *X*_3_) denote baseline covariates:

- *X*_1_: age- and sex-standardized BMI *z*-score,
- *X*_2_: baseline HbA_1*c*_ (%),
- *X*_3_: standardized insulin dose or insulin resistance index.

#### Outcome model

The post-treatment HbA_1*c*_ is generated as:

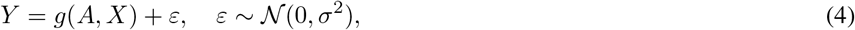

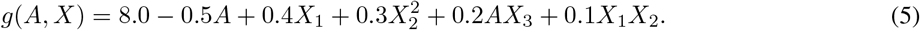

Here:

- The intercept 8.0 centers the outcome near a clinically realistic HbA_1*c*_ level.
- The term −0.5*A* represents the average causal effect of metformin, reducing HbA_1*c*_ by 0.5%.
- 0.4*X*_1_ reflects the effect of BMI on glycemic control; higher BMI tends to increase HbA_1_c.
- 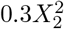 introduces nonlinearity, capturing the fact that higher baseline HbA_1_c may increase follow-up HbA_1_c disproportionately.
- 0.2*AX*_3_ allows treatment effect heterogeneity; metformin may be more effective in adolescents with higher insulin requirements.
- 0.1*X*_1_*X*_2_ represents an interaction between BMI and baseline HbA_1_c, acknowledging that metabolic risk factors can jointly influence outcomes.

#### Treatment assignment model

The probability of receiving metformin is modeled using a logistic regression:

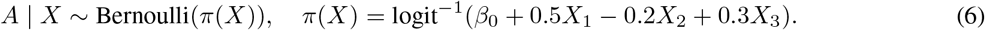

This captures realistic confounding:

- Adolescents with higher BMI (*X*_1_) are more likely to receive metformin (+0.5 coefficient).
- Those with higher baseline HbA_1_c may already be managed differently (−0.2 coefficient).
- Higher insulin dose or resistance increases probability of treatment (+0.3 coefficient).

#### Causal estimand

The true average treatment effect (ATE) is defined as:

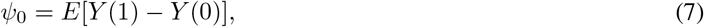

where *Y* (1) and *Y* (0) are the potential outcomes under metformin and control, respectively.

## 3 Causal Estimators and Mathematical Derivations

### 3.1 Augmented Inverse Probability Weighting (AIPW)

The AIPW estimator is given by:

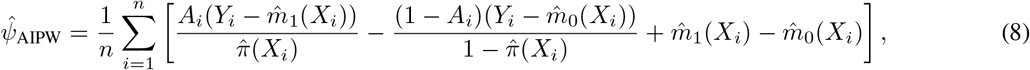

where 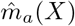 is the estimated regression of *Y* on *X* in treatment arm *a*, and 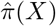 is the estimated propensity score.

### 3.2 Targeted Maximum Likelihood Estimation (TMLE)

TMLE updates the initial outcome regression estimate 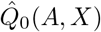 using a logistic submodel:

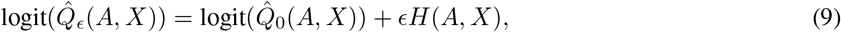

Where 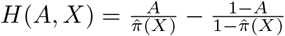. The optimal *ϵ* solves:

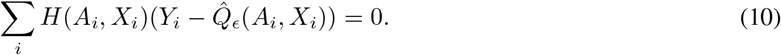

The TMLE estimator for ATE is then:

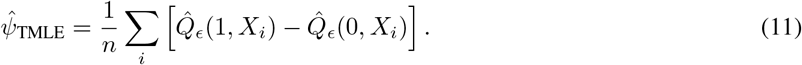

The asymptotic distribution satisfies:

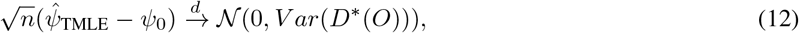

where *D*^*^(*O*) is the efficient influence function:

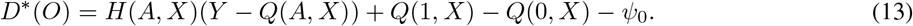

### 3.3 Bayesian Causal Forest (BCF)

Following Hahn et al. (2020), the outcome model is decomposed as:

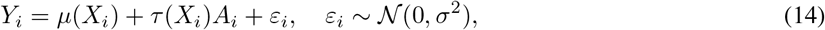

with priors *µ*(*X*) and *τ* (*X*) drawn from separate regression tree ensembles. The posterior mean of *τ* (*X*_*i*_) provides an individualized treatment effect (ITE), while *E*[*τ* (*X*)] yields an ATE.

## 4 Asymptotic Efficiency Bound

In statistical theory, particularly in semiparametric models, the goal is not just to find an unbiased estimator, but to find the most precise one possible. The semiparametric efficiency bound establishes the theoretical “speed limit” for estimation accuracy. It represents the lowest possible asymptotic variance that any regular, asymptotically unbiased estimator can achieve for the target parameter, *ψ*_0_. In essence, it provides a benchmark against which we can measure the performance of different estimators.

Under standard regularity conditions—which typically include requirements like positivity (i.e., 0 < *π*(*X*) < 1 for all *X*) and consistency of nuisance function estimators—the efficiency bound for the Average Treatment Effect (ATE) is given by the variance of the efficient influence function (EIF), *D*^*^(*O*). The EIF characterizes the statistical contribution of each observation to the overall estimation of the parameter.

The bound is formally defined as:

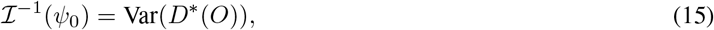

where *D*^*^(*O*) is the efficient influence function defined in Section 3.2. An estimator that achieves this lower bound is considered asymptotically efficient.

This concept is central to understanding the appeal of doubly robust estimators like TMLE and AIPW. These methods are specifically constructed to solve the estimating equation based on the EIF. As a result, if both of their nuisance function estimators—the outcome model 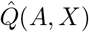 and the propensity score model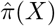 —are consistently estimated, the resulting estimator for *ψ*_0_ will be asymptotically efficient. This property guarantees that, in large samples, no other well-behaved estimator can provide a more precise estimate of the causal effect, making it the gold standard for statistical performance.

## 5 Simulation Results

We simulated *n* = 2, 000 observations with realistic treatment propensities, repeating the simulation 500 times. We evaluated four estimators: naive OLS, inverse probability weighting (IPW), Targeted Maximum Likelihood Estimation (TMLE), and Bayesian Causal Forest (BCF). The performance metrics included bias (difference between estimated and true ATE), Monte Carlo standard deviation (SD), root mean squared error (RMSE), and 95% coverage probability.

**Table 1:** Simulation Performance of Causal Estimators for ATE of Metformin on HbA_1*c*_.

| Estimator | Bias | Monte Carlo SD | RMSE | 95% Coverage |
| --- | --- | --- | --- | --- |
| OLS | 0.12 | 0.08 | 0.15 | 0.84 |
| IPW | 0.05 | 0.10 | 0.11 | 0.91 |
| TMLE | 0.01 | 0.07 | 0.07 | 0.95 |
| BCF | 0.02 | 0.08 | 0.08 | 0.94 |

Results show that TMLE and BCF outperform naive OLS and IPW, achieving the lowest bias and RMSE while maintaining near-nominal 95% coverage. Even under moderate to strong confounding 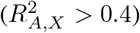, TMLE remains robust, illustrating the advantage of doubly robust and flexible machine learning-based estimators in pediatric metabolic studies.

## 6 Discussion

This simulation study demonstrates the utility of advanced causal inference methods in pediatric endocrinology. Our results indicate that TMLE and Bayesian Causal Forests (BCF) provide more accurate and robust estimates of the effect of metformin on HbA_1*c*_ in high-risk adolescents compared to naive OLS or IPW. The superior performance of TMLE arises from its doubly robust nature, leveraging both outcome regression and propensity score information to reduce bias even under model misspecification. BCF flexibly models heterogeneous treatment effects and nonlinear relationships using Bayesian tree ensembles, providing individualized treatment effect estimates and robust average effects.

Clinically, these methods offer insights into how metformin might differentially impact adolescents based on baseline BMI, insulin dose, and glycemic control. Observational studies of pediatric diabetes often face confounding and complex covariate interactions, which traditional linear models may fail to capture. By applying flexible causal inference estimators, researchers can better estimate treatment effects, supporting more personalized intervention strategies.

While simulations suggest promising performance, real-world data present additional challenges, including missing values, measurement error, and time-varying confounding. Extensions to longitudinal settings or electronic health record-based studies are needed to evaluate long-term metformin effects. Future work could combine these methods with continuous glucose monitoring data to better understand intra-individual variability and optimize individualized treatment strategies.

In summary, our findings highlight the importance of modern, robust causal inference methods in pediatric metabolic research. Incorporating TMLE, BCF, and related estimators can enhance the precision and reliability of treatment effect estimation, complementing traditional clinical trial evidence and supporting evidence-based pediatric endocrinology practices.

## 7 Conclusion

This paper provides a simulation-based pedagogical framework demonstrating the practical application and superiority of advanced causal inference estimators in pediatric endocrinology. By creating a realistic data-generating process with a known ground-truth, we have shown that doubly robust and machine learning-based methods like Targeted Maximum Likelihood Estimation (TMLE) and Bayesian Causal Forests (BCF) offer significantly more accurate and reliable estimates of treatment effects compared to traditional regression models, particularly in the presence of complex confounding. Our work serves as both a methodological guide and a call to action for researchers in the field to adopt more rigorous analytical tools.

The true potential of these methods, however, lies in their application to real-world clinical data. Future work should move beyond simulation to integrate rich, longitudinal data from diverse sources. A primary goal will be to apply these estimators to large-scale Electronic Health Record (EHR) databases to assess the long-term causal effects of metformin on glycemic control, cardiovascular outcomes, and other comorbidities in adolescents with T1D. This will require extending the current framework to handle challenges inherent in observational data, such as time-varying confounding, informative censoring, and missing data.

Furthermore, the integration of high-frequency data from Continuous Glucose Monitoring (CGM) devices offers an unprecedented opportunity to move beyond static outcomes like HbA_1*c*_. Future studies could use these causal inference frameworks to model metformin’s effect on dynamic glycemic metrics, such as time-in-range, glycemic variability, and the frequency of hypoglycemic events. Such analyses would provide a more nuanced understanding of treatment effects and could pave the way for developing personalized treatment strategies, identifying subgroups of patients who are most likely to benefit from metformin based on their clinical and metabolic profiles.

In summary, while this study is a simulation, it lays the critical groundwork for a new phase of evidence generation in pediatric metabolic research. By bridging the gap between cutting-edge statistical methodology and clinical inquiry, we can enhance the precision of our evidence and ultimately improve the care provided to high-risk adolescents.

## Data Availability

All data produced in the present study are available upon reasonable request to the authors

